# Plasma Neurofilament Light Chain and Incident Cardiovascular Outcomes in the General Population

**DOI:** 10.64898/2026.05.05.26352495

**Authors:** Jonah N. Keller, Jolie A. Kantor, Christopher Taveras, M Maria Glymour, Valentin Fuster, Fanny M. Elahi

**Author notes:** Corresponding Author. Fanny M. Elahi, MD, PhD. Contributed equally (Jolie A. Kantor, Christopher Taveras).

## Abstract

**Importance:** Standard cardiovascular risk scores fail to predict a substantial fraction of incident vascular clinical events in the general population. Whether circulating neurofilament light chain (NfL), a blood-based biomarker of neuronal axonal injury, independently predicts major cardiovascular or cerebrovascular disease outcomes in adults without known baseline clinical disease has not been tested.

**Objective:** To determine whether plasma NfL is independently associated with incident cardiac and brain vascular events in community-recruited adults and whether it adds prognostic information beyond established clinical risk factors.

**Design, Setting, and Participants:** Prospective cohort analysis within the UK Biobank. Plasma NfL was quantified using the Olink Explore proximity extension assay in 50,043 adults aged 40–69 years at recruitment. After excluding 3,914 participants with baseline known disease, the primary analysis included 46,129 adults. Incident events were ascertained via registry linkage over a median follow-up of 9.6 years.

**Exposures:** Plasma NfL, quantified in Olink Normalized Protein eXpression (NPX) units.

**Main Outcomes and Measures:** Primary outcome: major cardiovascular event (MVE), defined as the composite of cardiovascular death, nonfatal stroke, and nonfatal MI. Secondary outcomes: individual MVE components, heart failure hospitalization, and all-cause mortality.

**Results:** Among 46,129 participants free of baseline disease diagnoses, there were 2,912 incident MVE events. After adjustment for age, sex, BMI, smoking, hypertension, diabetes, and renal function, each one-NPX-unit increase in NfL was independently associated with MVE (aHR 1.46 (95% CI 1.36–1.57; *P* < .001)), nonfatal stroke (aHR 1.38 (95% CI 1.22–1.57; *P* < .001)), MI (aHR 1.35 (95% CI 1.22–1.50; *P* < .001)), cardiovascular death (aHR 1.79 (95% CI 1.59–2.01; *P* < .001)), heart failure hospitalization (aHR 1.50 (95% CI 1.36–1.65; *P* < .001)), and all-cause mortality (aHR 1.21 (95% CI 1.18–1.25; *P* < .001)). The associations were log-linear with no evidence of a threshold or saturation effect. Results were consistent in a sensitivity analysis including the full cohort with baseline vascular disease diagnoses. Addition of NfL to traditional risk factors improved 10-year discrimination most for cardiovascular death (C-statistic 0.809 to 0.817; ΔC = 0.007), heart failure (ΔC = 0.006), and stroke (ΔC = 0.008), with a smaller increment for the MVE composite.

**Conclusions and Relevance:** Plasma NfL was independently associated with incident cardiovascular and cerebrovascular outcomes among community-recruited adults after comprehensive risk-factor adjustments. These findings suggest that blood NfL levels may serve as a biomarker of subclinical vascular disease, strengthening early detection efforts and prompting further diagnostic evaluation.

## Introduction

A substantial fraction of incident cardiovascular events occurs in individuals who were previously asymptomatic or whose symptoms had been undetected and therefore untreated.^1–6^ This could in part be because healthcare systems engage in only limited screening for preclinical pathologies. In addition, traditional risk-factor thresholds may fail to risk stratify subclinical disease that precedes overt clinical events by years to decades.^7,8^ In addition, one of the most insidious forms of vascular disease, microvascular disease, can remain invisible to conventional risk-factor assessments, until larger blood vessels are affected and end-organ damage ensues.^9–11^ A potential readout of subclinical vascular pathology lies in the integrity and health of neuronal axons. Axons are particularly vulnerable to microvascular compromise and hypoperfusion.^12–16^ Accordingly, sensitive blood-based biomarkers of subclinical axonal injury, such as neurofilament light chain (NfL), may capture dimensions of vascular risk not reflected by conventional clinical measures, such as lipid profiles, blood pressure, and systemic inflammation.

NfL, a cytoskeletal protein released from injured axons, can be measured reliably and cost-effectively.^17,18^ NfL is a cytoskeletal protein of neuronal axons released into the bloodstream when axonal integrity is affected.^18–21^ Although NfL has historically been used to monitor neurological conditions such as multiple sclerosis and Alzheimer disease,^17–19,22–25^ accumulating evidence indicates that it is also elevated in peripheral vascular disease, cerebrovascular conditions, as well as systemic contexts such as peripheral neuropathies, and kidney disease.^26–31^ In patients with suspected coronary artery disease, baseline NfL predicted cardiovascular death with an AUC of 0.85 at one year.^32^ In heart failure with reduced ejection fraction, NfL rivalled NT-proBNP in mortality discrimination (C-index 0.70).^33^ Cross-sectional NHANES data showed NfL to be associated with prevalent cardiovascular disease, with strong associations among non-hypertensive adults (adjusted OR 2.72; 95% CI 1.61–4.62).^34^ These observations raise the question of whether NfL may be independently associated with future cardiovascular and cerebrovascular disease clinical outcomes independent of vascular risk factors.

While cross-sectional associations with prevalent disease are abundant, prospective evidence for the predictive value of NfL for vascular outcomes related to the heart and brain remains limited. Prior studies have been conducted predominantly in disease-enriched cohorts, including patients with established coronary disease, heart failure, atrial fibrillation, and cerebrovascular disease.^32,33,35^ This leaves the question unanswered of whether NfL can captured subclinical vascular disease and predict future incident vascular events when measured in adults without overt cardiac or cerebrovascular diseases.

Herein, we describe a study using the UK Biobank (UKB), to evaluate NfL as an independent, predictive blood-based biomarker for clinically significant vascular outcomes of heart and brain in community-recruited adults.

## Methods

### Participants

The UK Biobank is a prospective cohort that recruited 503,325 adults aged 40–69 years at 22 assessment centers across England, Scotland, and Wales between 2006 and 2010.^36^ At baseline, participants completed detailed questionnaires, underwent anthropometric and clinical measurements, and provided blood samples for long-term storage. Ethics approval was granted by the North West Multi-Centre Research Ethics Committee (ref. 06/MRE08/65), and all participants provided written informed consent.

The present analysis includes 50,043 participants with available NfL measurements from the UKB Olink Explore proteomics release (application 21234) and complete covariate data. Because the primary objective was to evaluate NfL as a screening biomarker for individuals without established cardiovascular or cerebrovascular diseases, participants with known clinically overt disease at enrolment were excluded: coronary artery disease (2,181; 4.4%), prior stroke or TIA (1,151; 2.3%), atrial fibrillation (1,012; 2%), heart failure (426; 0.9%), and peripheral artery disease (164; 0.3%). After exclusion of 3,914 participants with known vascular diseases (7.8%), the primary analysis cohort comprised 46,129 adults free of heart and brain clinical vascular disease diagnoses at baseline. A sensitivity analysis including the full cohort of 50,043 participants (with prevalent vascular diseases) was also performed.

### Biomarker Assay

Plasma NfL was quantified using the Olink Explore 1536 proximity extension assay (PEA; Olink Proteomics AB, Uppsala, Sweden), which targets the NEFL gene product (neurofilament light polypeptide; UniProt P07196). Values are reported in Normalized Protein eXpression (NPX) units, a relative scale derived from log_2_-transformed next-generation sequencing counts that is normalized across samples and plates; higher values indicate higher relative protein abundance. NPX values are centered near zero and may be negative. No additional log-transformation was applied. We refer to this measurement as NfL throughout.

### Outcomes

Outcomes were ascertained via linkage to Hospital Episode Statistics (HES; ICD-10) and the Office for National Statistics (ONS) death registry. The primary outcome was major vascular event (MVE), defined as a composite of three hard endpoints: cardiovascular death, nonfatal stroke, and nonfatal myocardial infarction. Nonfatal stroke was defined as ischemic stroke (ICD-10 I63) or stroke not specified as hemorrhagic or ischemic (I64) recorded as the primary HES diagnosis; intracerebral hemorrhage (I61) was not included. Nonfatal myocardial infarction was identified by ICD-10 I21–I22 in HES or the UKB algorithmic definition (field 42000). Secondary outcomes comprised each component of the MVE composite analyzed individually, plus two additional endpoints: heart failure hospitalization (ICD-10 I50 as primary or secondary HES diagnosis with at least one overnight stay) and all-cause mortality. Cardiovascular death was defined as death with an underlying cause classified to ICD-10 chapter I in the ONS death registry.

### Statistical Analysis

Participant characteristics were tabulated by NfL quartile. Three Cox proportional hazards models were fitted for each outcome: unadjusted; age- and sex-adjusted; and multivariable-adjusted for age, sex, BMI, smoking status, hypertension, diabetes mellitus, and eGFR (CKD-EPI). The functional form of the NfL–log-hazard relationship was evaluated with restricted cubic splines (three knots at the 10th, 50th, and 90th percentiles); spline curves were consistent with approximate linearity for all outcomes. Pre-specified subgroup analyses examined effect modification by sex and baseline age group (< 60 vs ≥ 60 years) using multiplicative interaction terms. Incremental prognostic value was quantified as the change in Harrell’s C-statistic. A pre-specified sensitivity analysis repeated all Cox and C-statistic analyses in the full cohort (including participants with prevalent cardiovascular disease at baseline), with additional adjustment for coronary artery disease, prior stroke or TIA, heart failure, and peripheral artery disease. All analyses were performed in R version 4.4 (R Foundation for Statistical Computing).

## Results

### Cohort characteristics

After exclusion of 3,914 participants with prevalent cardiovascular disease, the primary analysis cohort comprised 46,129 adults free of cardiovascular disease at baseline (mean age 56.4 ± 8.2 years; 44% male) with a median follow-up of 9.6 years (**Table 1**).

**Table 1.** Baseline characteristics of the primary prevention cohort by NfL quartile.

| Characteristic | Overall | 1 (lowest NfL) | 2 | 3 | 4 (highest NfL) | P for trend |
| --- | --- | --- | --- | --- | --- | --- |
| n | 46129 | 11533 | 11532 | 11532 | 11532 |  |
| Age, years (mean (SD)) | 56.38 (8.21) | 50.37 (7.27) | 54.89 (7.56) | 58.51 (7.04) | 61.74 (6.21) | <0.001 |
| Male sex, n | 20430 (44.3) | 5105 (44.3) | 5070 (44.0) | 5102 (44.2) | 5153 (44.7) | 0.027 |
| BMI, kg/m<sup>2</sup> (mean (SD)) | 27.32 (4.74) | 28.06 (5.19) | 27.42 (4.69) | 27.10 (4.51) | 26.70 (4.45) | <0.001 |
| Smoking status |  |  |  |  |  | <0.001 |
| Current | 4836 (10.5) | 1369 (11.9) | 1302 (11.3) | 1125 (9.8) | 1040 (9.1) |  |
| Never | 25568 (55.7) | 6600 (57.5) | 6430 (56.0) | 6330 (55.2) | 6208 (54.1) |  |
| Previous | 15512 (33.8) | 3515 (30.6) | 3747 (32.6) | 4017 (35.0) | 4233 (36.9) |  |
| eGFR, mL/min/1.73m<sup>2</sup> (median [IQR]) | 97.34 [87.20, 103.87] | 102.85 [94.50, 108.89] | 98.78 [89.92, 104.83] | 95.98 [85.86, 101.64] | 92.14 [79.78, 98.78] | <0.001 |
| CRP, mg/L (median [IQR]) | 1.32 [0.66, 2.75] | 1.28 [0.62, 2.76] | 1.29 [0.64, 2.68] | 1.32 [0.66, 2.72] | 1.40 [0.70, 2.88] | <0.001 |
| NfL, NPX (median [IQR]) | -0.02 [-0.37, 0.33] | -0.61 [-0.81, -0.48] | -0.18 [-0.27, -0.10] | 0.14 [0.05, 0.23] | 0.59 [0.45, 0.82] | <0.001 |
| Hypertension, n | 10897 (23.6) | 2144 (18.6) | 2460 (21.3) | 2782 (24.1) | 3511 (30.4) | <0.001 |
| Diabetes mellitus, n | 2009 (4.4) | 376 (3.3) | 404 (3.5) | 491 (4.3) | 738 (6.4) | <0.001 |

### NfL distribution and event rates

Event incidence increased monotonically with NfL quartile for every outcome, from 0.44 (0.40-0.48) to 1.30 (1.22-1.37) MVE events per 100 person-years from Q1 to Q4 (**Table 2**). Kaplan– Meier curves for the primary composite (major vascular event) showed early and progressive separation across NfL quartiles that widened continuously throughout follow-up, with 15-year cumulative incidence of 6.9% in Q1 versus 19.4% in Q4 (log-rank *P* <.001; **Figure 1A**).

**Table 2.** Events and incidence rates per 100 person-years (95% CI) by NfL quartile, with Q4/Q1 incidence rate ratio and log-rank P.

| Outcome | Q1 | Q2 | Q3 | Q4 | Total | Q4/Q1<br>Rate<br>Ratio | Log-rank P |
| --- | --- | --- | --- | --- | --- | --- | --- |
| Major Vascular Event | 436<br>0.44 (0.40-0.48) | 610<br>0.61<br>(0.56-0.66) | 709<br>0.73<br>(0.68-0.79) | 1157<br>1.30<br>(1.22-1.37) | 2912<br>0.76<br>(0.73-0.78) | 3.0 | <.001 |
| Nonfatal Stroke | 144<br>0.14 (0.12-0.17) | 214<br>0.21<br>(0.18-0.24) | 223<br>0.23<br>(0.20-0.26) | 362<br>0.40<br>(0.36-0.44) | 943<br>0.24<br>(0.23-0.26) | 2.8 | <.001 |
| Nonfatal MI | 227<br>0.23 (0.20-0.26) | 304<br>0.30<br>(0.27-0.34) | 359<br>0.37<br>(0.33-0.41) | 524<br>0.58<br>(0.53-0.63) | 1414<br>0.36<br>(0.34-0.38) | 2.6 | <.001 |
| CV Death | 87<br>0.09 (0.07-0.11) | 125<br>0.13<br>(0.10-0.15) | 160<br>0.16<br>(0.14-0.19) | 364<br>0.41<br>(0.36-0.45) | 736<br>0.19<br>(0.18-0.20) | 4.7 | <.001 |
| HF Hospitalization | 203<br>0.20 (0.17-0.23) | 317<br>0.32<br>(0.28-0.35) | 372<br>0.38<br>(0.34-0.42) | 652<br>0.72<br>(0.66-0.77) | 1544<br>0.40<br>(0.38-0.42) | 3.6 | <.001 |
| All-Cause Mortality | 2929<br>2.93 (2.82-3.03) | 3644<br>3.65<br>(3.53-3.77) | 4333<br>4.47<br>(4.33-4.60) | 5436<br>6.06<br>(5.90-6.22) | 16342<br>4.23<br>(4.16-4.29) | 2.1 | <.001 |

**Figure 1.**
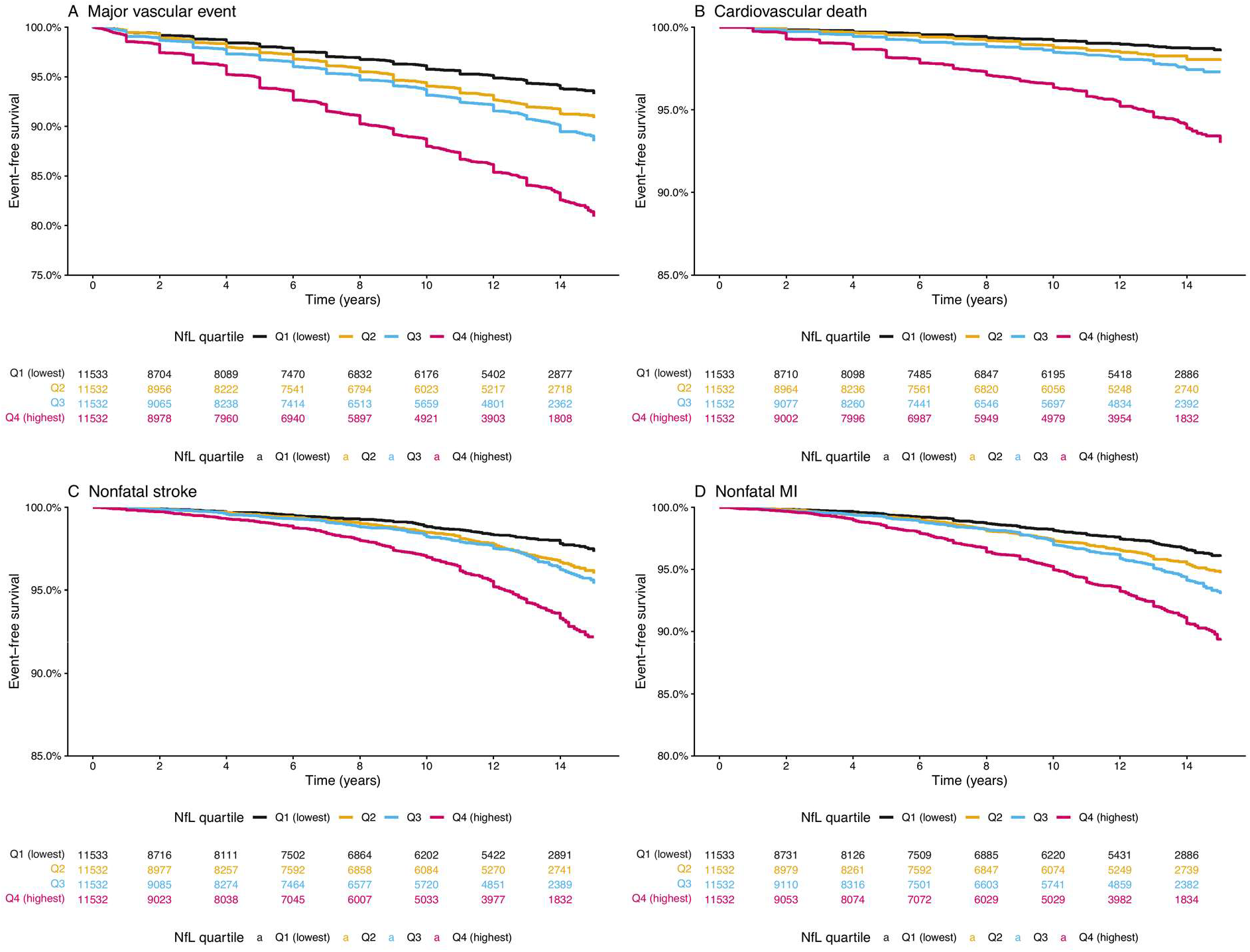
Kaplan–Meier event-free survival by NfL quartile. (1A) Major vascular event (primary composite). (1B) Cardiovascular death. (1C) Nonfatal stroke. (1D) Nonfatal myocardial infarction. Follow-up truncated at 15 years. Number at risk shown below each panel. Log-rank P < .001 for all comparisons. Q1 = lowest NfL quartile; Q4 = highest.

Cardiovascular death showed the most pronounced quartile separation (Q1 1.4% vs Q4 7.0%; log-rank *P* <.001), with a clear dose–response gradient that was apparent within the first two years of follow-up (**Figure 1B**). Nonfatal stroke showed a similar pattern with progressive divergence across quartiles (Q1 2.8% vs Q4 8.3%; log-rank *P* <.001; **Figure 1C**). The gradient was also present for nonfatal MI (Q1 4.1% vs Q4 11.2%; log-rank *P* <.001), though the separation between the intermediate quartiles (Q2, Q3) was less distinct (**Figure 1D**). All log-rank *P* values were <.001.

### Association with outcomes

After multivariable adjustment, each one-NPX-unit increase in NfL was associated with MVE (aHR 1.46 (95% CI 1.36–1.57; *P* < .001)), nonfatal stroke (aHR 1.38 (95% CI 1.22–1.57; *P* < .001)), nonfatal MI (aHR 1.35 (95% CI 1.22–1.50; *P* < .001)), cardiovascular death (aHR 1.79 (95% CI 1.59–2.01; *P* < .001)), heart failure hospitalization (aHR 1.50 (95% CI 1.36–1.65; *P* < .001)), and all-cause mortality (aHR 1.21 (95% CI 1.18–1.25; *P* < .001)) (**Table 3**). Attenuation from unadjusted to multivariable estimates was statistically significant for all outcomes (38– 61%; all Freedman–Schatzkin *P* < 0.001), indicating that age and sex explain a substantial but incomplete proportion of the NfL–outcome association; HRs were almost identical after additional adjustment for other risk factors. Restricted cubic spline analyses confirmed that the dose–response relationship was log-linear for all six outcomes, with no evidence of a threshold below which NfL lacked prognostic value and no saturation at high concentrations (**eFigure 1**). The spline curves for cardiovascular death and heart failure hospitalization showed the steepest gradients, while the curves for nonfatal MI and all-cause mortality were flatter but remained monotonically increasing across the NfL distribution.

**Table 3.** Hazard ratio (95% CI) per 1-NPX-unit increase in NfL for each cardiovascular outcome, by model. Multivariable model adjusted for age, sex, BMI, eGFR, smoking, hypertension, and diabetes mellitus. Primary prevention cohort (prevalent CVD excluded).

| Outcome | Unadjusted | Age & sex adjusted | Multivariable | % Attenuation | P attenuation |
| --- | --- | --- | --- | --- | --- |
| Major Vascular Event | 2.07 (1.96–2.18) | 1.48 (1.38–1.59) | 1.46 (1.36–1.57) | 48% | <0.001 |
| Nonfatal Stroke | 2.09 (1.91–2.30) | 1.42 (1.25–1.61) | 1.38 (1.22–1.57) | 56% | <0.001 |
| Nonfatal MI | 1.93 (1.78–2.08) | 1.35 (1.22–1.49) | 1.35 (1.22–1.50) | 54% | <0.001 |
| CV Death | 2.54 (2.33–2.76) | 1.91 (1.70–2.14) | 1.79 (1.59–2.01) | 38% | <0.001 |
| HF Hospitalization | 2.28 (2.13–2.43) | 1.49 (1.35–1.64) | 1.50 (1.36–1.65) | 51% | <0.001 |
| All-Cause Mortality | 1.65 (1.61–1.70) | 1.18 (1.14–1.21) | 1.21 (1.18–1.25) | 61% | <0.001 |

### Subgroup and interaction analyses

In a sensitivity analysis including the full cohort of 50,043 participants (including 3,914 with prevalent cardiovascular disease at enrolment), NfL remained independently associated with incident MVE after additional adjustment for prevalent cardiovascular conditions (aHR 1.40 (95% CI 1.32–1.49; *P* <.001)). C-statistics in the full cohort were similar to the primary analysis (**eTable 2**).

Subgroup analyses for the primary composite endpoint (major vascular event) showed statistically significant effect modification by sex and a borderline interaction with the age dichotomy (**Figure 2**). The association was nominally stronger in participants aged under 60 (aHR 1.66 (95% CI 1.49–1.84)) than in those aged 60 or older (aHR 1.48 (95% CI 1.36–1.61); *P*_int_ = 0.051). Hazard ratios were higher in women (aHR 1.51 (95% CI 1.34–1.70)) than in men (aHR 1.44 (95% CI 1.32–1.56); *P*_int_ = 0.004). In both cases, the direction of association was consistent across strata; the interaction reflected differences in magnitude rather than direction.

**Figure 2.**
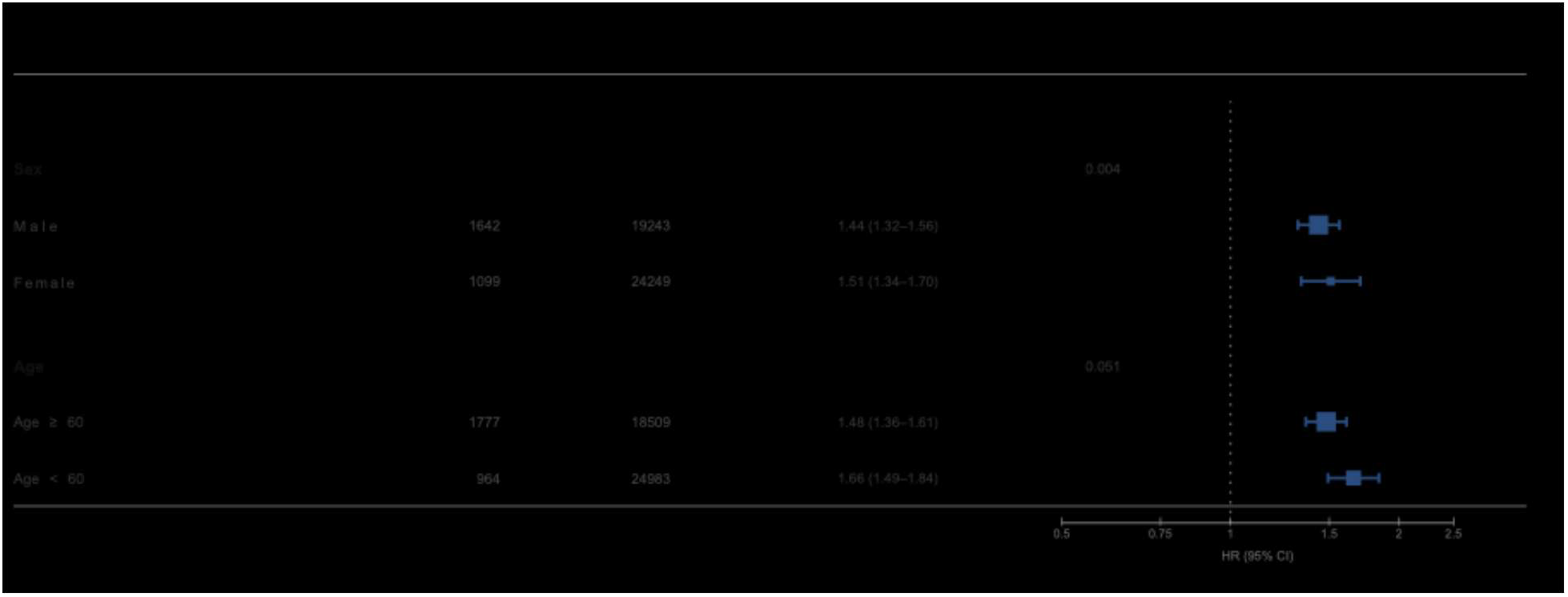
Subgroup analyses for the primary composite endpoint (major vascular event) by age group and sex. Hazard ratios per 1-NPX-unit increase in NfL from multivariable Cox models within each stratum. Squares indicate point estimates; horizontal bars indicate 95% confidence intervals. Interaction P values from likelihood-ratio testing.

To further characterize the NfL–hazard relationship by sex, dose–response curves were plotted separately for men and women across all six outcomes (**eFigure 2**). For the primary composite (major vascular event), male and female curves were broadly overlapping despite the statistically significant categorical interaction, suggesting that the difference in magnitude is modest on the continuous scale. The curves for cardiovascular death and heart failure hospitalization showed similarly concordant patterns between sexes. For nonfatal stroke, a modest divergence was apparent at higher NfL concentrations, with a steeper gradient in men, though confidence intervals overlapped.

Female and male spline curves diverged significantly for the primary MVE endpoint (NfL × sex interaction *P* = 0.011), indicating sex to be a meaningful variable and the importance of sex-stratified risk estimates in future clinical implementation of these findings.

Age-stratified splines were fitted independently within three 10-year age bands for the MVE endpoint to characterize the NfL–hazard relationship within each age group (**eFigure 3**). These curves depict the within-group association between NfL and outcome and are not intended for direct comparison of absolute risk across age categories.

With stratification of the cohort into three age bands, the NfL–MVE dose–response curve was steepest in younger participants (< 55 years) in comparison to those aged 65 or older. This finding did not reach statistical significance, with overlapping confidence intervals (**eFigure 3**). The continuous NfL × age interaction test yielded *P* = 0.133. Age-specific dose–response curves evaluated at six ages from 45 to 70 years confirmed that the NfL–MVE relationship was present across the full age range, with the steepest gradient at younger ages and a progressively flatter slope in older participants (**eFigure 4**).

The age-dependent NfL–MVE relationship did not differ significantly between men and women; male and female hazard trajectories were broadly parallel across the recruitment age range, with no significant three-way sex × age × NfL interaction (**eFigure 5**).

### Incremental discrimination and risk stratification

Addition of NfL to models containing traditional vascular risk factors was associated with statistically significant improvements in discrimination across multiple outcomes, including cardiovascular death, stroke, and heart failure **(eTable 1)**. In the primary prevention cohort, NfL modestly improved prediction of 10-year major vascular events when added to clinical risk models, with consistent findings across alternative model specifications **(eTable 3)**. These patterns were similar across age and sex strata **(eTable 4**). Stratified analyses combining age, sex, and NfL quartiles demonstrated a broad gradient in observed 10-year risk across subgroups **(eFigure 6)**.

## Discussion

### Principal findings

At present, risk stratification for cardiovascular and cerebrovascular disease is costly and rarely implemented at a population scale to detect preclinical disease prior to tissue injury.^37,38^ This gap is consequential, as a substantial proportion of events occur in individuals classified as low or intermediate risk by established scores. An estimated 41% of cardiovascular events occur in individuals deemed low risk, and 57% of ASCVD events occur in those below the statin-eligibility threshold, underscoring the need for cost-effective biomarkers that can identify risk below current action thresholds.^1,39–43^ In 46,129 community-recruited adults free of cardiovascular disease at baseline, a single measurement of plasma NfL was independently associated with incident major vascular events, nonfatal stroke, nonfatal MI, cardiovascular death, heart failure hospitalization, and all-cause mortality over a median of 9.6 years. The association was graded, log-linear, and robust to multivariable adjustment, and was not attenuated when the full cohort including prevalent cardiovascular disease was analyzed. Neuronal axons are exquisitely sensitive to vascular injury. Accordingly, circulating levels of NfL can capture subclinical disease well before overt end-organ damage manifests in the heart or brain. Our findings extend prior NfL–vascular associations from disease-enriched cohorts to a primary prevention population, and suggest that plasma NfL may capture subclinical vascular risk invisible to conventional risk-factor assessment.^17,18,33,35^ As such levels of NfL can capture subclinical disease well before frank end organ damage can occur affecting the heart and the brain.

### Comparison with prior studies

Prior evidence linking NfL to cardiovascular and cerebrovascular outcomes has come predominantly from patients with established clinical disease. In suspected coronary artery disease, baseline NfL discriminated cardiovascular death with an AUC of 0.85 at one year, and in heart failure with reduced ejection fraction its prognostic accuracy rivalled that of NT-proBNP (C-index 0.70).^32,33^ Among patients with atrial fibrillation, each doubling of NfL was associated with a 35% higher risk of major vascular events.^35^ Cross-sectional NHANES data extended these observations to community-dwelling adults, with particularly strong associations among those without hypertension (OR 2.72; 95% CI 1.61–4.62). ^34^ Taken together, these studies have established the biological plausibility and prognostic relevance of circulating NfL in cardiovascular disease. However, all prior studies were conducted in disease-enriched or cross-sectional settings, leaving unanswered whether the signal holds prospectively in an unselected population without prevalent cardiac disease. The present analysis addresses this gap directly, and the consistency of the association across six prespecified hard endpoints, together with the full-cohort sensitivity analysis, provides evidence that the NfL– cardio/cerebrovascular relationship is not an artefact of reverse causation or prevalent disease confounding.

### Biological interpretation of NfL as a predictive biomarker for cardiovascular disease

The mechanism linking a marker of neuro-axonal injury to cardiovascular events is not established, but several pathways are plausible. Subclinical cerebrovascular disease, chronic small vessel disease, and silent microinfarcts and lacunar strokes, are common in middle-aged adults without clinically recognized cardiovascular or cerebrovascular disease.^44–48^ Neuronal axons, the source of NfL levels, are highly sensitive to microvascular dysfunction and hypoperfusion.^12–14,21^ Because these vascular pathologies share upstream risk pathways with coronary and peripheral artery disease, elevated NfL may reflect shared small-vessel disease related injury that has affected neurons in the periphery or central nervous system before it is clinically apparent in the heart.^49–51^ Cardiac dysfunction can itself promote neuroinflammation and axonal injury through circulating pro-inflammatory mediators, and shared drivers such as hypertension, hyperglycemia, age-related immune dysfunction and sterile inflammation, all can contribute to neuronal axonal injury.^52–56^ In addition, abnormal clearance of NfL, related to chronic kidney disease, another manifestation of small vessel disease, can also be independent of primary neurological disease, cause elevation in NfL.^49,57^ The present data cannot distinguish among these mechanisms. The observation that NfL adds prognostic information after adjustment for renal function, inflammation, hypertension, and metabolic comorbidities suggests it may capture dimensions of vascular injury not reflected by these conventional risk factors alone. This residual signal raises a broader question: whether individual vulnerability to vascular risk factors is sufficiently heterogeneous that population-based thresholds are inadequate for meaningful optimization of heart and brain healthspan.

### Limitations

Several limitations warrant consideration. First, the UK Biobank is subject to volunteer selection bias, which attenuates event rates and may compress effect sizes.^36^ Second, NfL was measured at a single time point; serial measurements may better capture trajectories of neurovascular risk than a single baseline value. Third, residual confounding from unmeasured variables, including subclinical small vessel disease reflected by neuroimaging or cardiovascular imaging, detailed lipid subfractions, and unrecorded atrial fibrillation, cannot be excluded. Given the goal of assessing NfL as a screening tool, we did not leverage any imaging variables. Finally, the Olink proximity extension assay provides relative (NPX) rather than absolute concentrations, and per-unit hazard ratios are not directly interchangeable with those from clinical immunoassay platforms such as Simoa (Quanterix).

### Implications

The central finding of this study is that a marker of neuro-axonal injury independently predicts cardiovascular events in a population without known cardiac disease. This suggests that subclinical neurovascular damage and cardiovascular risk share upstream biology that is not adequately captured by current screening approaches, and that the nervous system may serve as an early sentinel of vascular compromise detectable in blood before clinical cardiac disease manifests. The strongest improvements in discrimination were observed for cardiovascular death, heart failure, and stroke, the endpoints most plausibly linked to microvascular and neurovascular pathophysiology. If validated, NfL could offer a scalable, blood-based complement to existing risk algorithms, particularly for identifying individuals at low-to-intermediate predicted risk who harbor subclinical vascular disease that would otherwise go undetected. Incorporation of NfL into clinical risk models resulted in statistically significant improvements in risk discrimination across multiple cardiovascular outcomes and enabled meaningful stratification across a broad range of absolute risk, supporting its potential role in refining current risk assessment strategies.

## Conclusions

In 46,129 community-recruited adults free of cardiovascular disease at baseline, plasma NfL was independently associated with incident cardiovascular and cerebrovascular events across a graded dose–response and after comprehensive risk-factor adjustment. The association was consistent in a sensitivity analysis including the full cohort with prevalent cardiovascular disease and persisted over up to 15 years of follow-up. The consistency of this signal across multiple endpoints and demographic strata, together with its biological plausibility, supports further evaluation of NfL as a potential sensitive biomarker of vascular disease with implications for risk stratification and preclinical intervention of individuals to prevent cardiovascular and cerebrovascular outcomes. Future studies should test the value of NfL prospectively to determine whether it can become a clinically actionable risk stratifier for brain and heart vascular outcomes. If NfL can detect and stratify subclinical vascular disease, the implications for healthspan-focused interventions are substantial.

## Data Availability

The data that support the findings of this study are available from the UK Biobank. Restrictions apply to the availability of these data, which were used under application number 21234, and so are not publicly available. Data are, however, available from the UK Biobank upon reasonable request and with appropriate approvals (www.ukbiobank.ac.uk).

## Acknowledgements

This research was conducted using the UK Biobank resource under application number 21234. This research was supported by the Edward and Pearl Fein gift to F.M.E.; the National Institute on Aging and Department of Veterans Affairs IK2CX002180, Larry L. Hillblom Foundation 2019A012SUP and New Vision Research to F.M.E; the Dolby Family Fund, the Simon Family Trust, Brightfocus, NIH/NIA RF1 AG064926, and NIH/NINDS R35 NS097976; NIH R01NS122888, UH3NS106899, U24NS122732, US VA 1I01RX002245, I01RX002787, I01BX005871, I50BX005878, Craig H. Neilsen Foundation, and Wings for Life Foundation. We thank the Gladstone Bioinformatics Core for their interactive enrichment analysis tools.

## Author contributions

FME conceived the study. JNK and FME designed the detailed analytical plan. JNK performed the biostatistical analyses and generated figures. JNK, MMG, VF, and FME critically discussed and interpreted the results. JNK, JAK, CT, MMG, VF, and FME drafted the manuscript. All authors critically read and provided edits. FME provided funding.

